# The inactivated NDV-HXP-S COVID-19 vaccine induces a significantly higher ratio of neutralizing to non-neutralizing antibodies in humans as compared to mRNA vaccines

**DOI:** 10.1101/2022.01.25.22269808

**Authors:** Juan Manuel Carreño, Ariel Raskin, Gagandeep Singh, Johnstone Tcheou, Hisaaki Kawabata, Charles Gleason, Komal Srivastava, Vladimir Vigdorovich, Nicholas Dambrauskas, Sneh Lata Gupta, Irene Gonzalez, Jose Luis Martinez, Stefan Slamanig, D. Noah Sather, Rama Raghunandan, Ponthip Wirachwong, Sant Muangnoicharoen, Punnee Pitisuttithum, Jens Wrammert, Mehul S. Suthar, Weina Sun, Peter Palese, Adolfo García-Sastre, Viviana Simon, Florian Krammer

## Abstract

NDV-HXP-S is a recombinant Newcastle disease virus based-vaccine against severe acute respiratory syndrome coronavirus 2 (SARS-CoV-2), which expresses an optimized (HexaPro) spike protein on its surface. The vaccine can be produced in embryonated chicken eggs using the same process as that employed for the production of influenza virus vaccines. Here we performed a secondary analysis of the antibody responses after vaccination with inactivated NDV-HXP-S in a Phase I clinical study in Thailand.

The SARS-CoV-2 neutralizing and spike binding activity of NDV-HXP-S post-vaccination serum samples was compared to that of matched samples from mRNA BNT162b2 (Pfizer) vaccinees. Neutralizing activity of sera from NDV-HXP-S vaccinees was comparable to that of individuals vaccinated with BNT162b2. Interstingly, the spike binding activity of the NDV-HXP-S vaccinee samples was lower than that of sera obtained from individuals vaccinated with the mRNA vaccine. This let us to calculate ratios between binding and neutralizing antibody titers. Samples from NDV-HXP-S vaccinees had binding to neutralizing activity ratios similar to those of convalescent sera suggesting a very high proportion of neutralizing antibodies and low non-neutralizing antibody titers. Further analysis showed that, in contrast to mRNA vaccination, which induces strong antibody titers to the receptor binding domain (RBD), the N-terminal domain, and the S2 domain, NDV-HXP-S vaccination induces a very RBD focused response with little reactivity to S2. This explains the high proportion of neutralizing antibodies since most neutralizing epitopes are located in the RBD. In conclusion, vaccination with inactivated NDV-HXP-S induces a high proportion of neutralizing antibodies and absolute neutralizing antibody titers comparable to those after mRNA vaccination.

## Introduction

A large number of vaccines for severe acute respiratory syndrome coronavirus 2 (SARS-CoV-2) have been developed and licensed (*1*). Nevertheless, there is a need for SARS-CoV-2 vaccines that can be produced at low cost locally in low- and middle-income countries (LMICs). The NDV-HXP-S vaccine (*2*) is based on a Newcastle disease virus (NDV) vector which presents a stabilized HexaPro (*3*) version of the spike protein on its surface. This vaccine can be manufactured like influenza virus vaccines at low cost in embryonated chicken eggs in facilities located globally, including in LMICs (*2, 4-6*). NDV-HXP-S can be used as live vaccine (*2, 7, 8*) or as an inactivated vaccine (*2, 9*). Clinical trials with a live version are ongoing in Mexico (NCT04871737) and the US (NCT05181709), while the inactivated vaccine is being tested in Vietnam (NCT04830800), Thailand (NCT04764422,) and Brazil (NCT04993209). Results from the initial Phase I trials are promising and Phase I data from Thailand with the inactivated vaccine have been reported (*9*). Phase II trials with the inactivated vaccine have also been successfully conducted, while Phase III trials are currently in the planning stage.

It has been shown that both natural infection- and vaccine-induced immunity target different parts of the SARS-CoV-2 spike protein, including the receptor binding domain (RBD), the N-terminal domain (NTD), and the S2 domain (*10-15*). Most described neutralizing epitopes can be found on the RBD and the NTD, while very few S2 directed antibodies neutralize the virus *in vitro* (*12, 16, 17*). Furthermore, it has been shown that the ratio of neutralizing to non-neutralizing antibodies differs between natural infection and mRNA vaccination (*10, 15*). While mRNA vaccination induces higher absolute neutralizing antibody titers in serum, infection induces a higher proportion of neutralizing antibodies. In other words, a large percentage of mRNA vaccine-induced antibodies bind spike but do not neutralize the virus, while this percentage is lower after natural infection. In addition, it is known that, while neutralizing activity can be drastically reduced against viral variants, binding activity is better retained (*18, 19*). In this case, of course, the ratio between neutralizing and non-neutralizing antibodies also changes. While both neutralizing and binding antibodies have been implicated as correlates of protection (*20*), only neutralizing antibodies are likely to block infection.

Here we performed a secondary analysis comparing sera from individuals vaccinated with NDV-HXP-S in Thailand to sera from convalescent and mRNA vaccinated individuals collected under observational cohort studies in New York City (e.g., PARIS study (*21*)) to investigate neutralizing activity, ratios of binding to neutralizing antibodies and activity against variants of concern.

## Results

### Serum samples from vaccinees and convalescent individuals

Two sets of sera where used for this study. The first set comprised of sera from a clinical trial in Thailand (NCT04764422) (*9*) which included six groups: a placebo control group (n=35), a group that received 1μg of inactivated NDV-HXP-S (n=35), a group that received 1μg plus ODN1018 adjuvant (n=35), a group that received 3μg (n=35), a group that received 3 μg plus ODN1018 (n=35) and a group that received 10μg of NDV-HXP-S (n=35). Individuals were vaccinated twice, on day 1 and day 29; sera tested were collected two weeks after the boost (day 43). The second set of sera were from observational longitudinal studies conducted in New York City and comprised of serum samples from 20 study participants (PARIS) who received the BNT162b2 (Pfizer) mRNA vaccine and 18 serum samples from convalescent individuals (infected with prototype SARS-CoV-2; PARIS as well as viral infection cohorts). Sera were collected approximately 14 days post 2^nd^ dose for the BNT162b2 vaccinees and approximately four weeks post infection for convalescent individuals. Age ranges and sex distribution between the samples from Thailand and New York were comparable (**Table 1**).

**Table 1:** Sample characteristics.

|  | Age<sup>1</sup> (range) | Sex | Days post exposure (range or protocol specified maximum deviation) |
| --- | --- | --- | --- |
| Placebo | 33 (26-39) | 60% female | 14 (11-17)<sup>2</sup> |
| 1 ug NDV-HXP-S | 39 (32-45) | 60% female | 14 (11-17)<sup>2</sup> |
| 1 ug NDV-HXP-S + ODN1018 | 37 (29-49) | 80% female | 14 (11-17)<sup>2</sup> |
| 3 ug NDV-HXP-S | 34 (25-44) | 57.1 % female | 14 (11-17)<sup>2</sup> |
| 3 ug NDV-HXP-S + ODN1018 | 37 (31-42) | 48.6 % female | 14 (11-17)<sup>2</sup> |
| 10 ug NDV-HXP-S | 32 (27-42) | 60% female | 14 (11-17)<sup>2</sup> |
| BNT162b2 (Pfizer) | 39 (23-48) | 80% female | 15 (10-20)<sup>3</sup> |
| Healthy convalescent | 39 (26-59) | 55.6% female | 29.5 (14-49)<sup>4</sup> |

<sup>1</sup>median
<sup>2</sup>post 2<sup>nd</sup> dose, target interval plus maximum protocol deviation in parenthesis
<sup>3</sup>post 2<sup>nd</sup> dose for mRNA vaccinees, median plus range
<sup>4</sup>post-PCR confirmation for convalescent individuals , median plus range

### Neutralizing activity against wild type SARS-CoV-2 of sera from NDV-HXP-S vaccinees is similar to that of sera from BNT162b2 vaccinees

First, we tested the neutralizing activity of the sera from the NDV-HXP-S trial (placebo, 1μg, 1 μg + ODN1018, 3μg, 3μg + ODN1018, and 10μg) as well as the BNT162b2 and the human convalescent sera (HCS) against wild type SARS-CoV-2. Few individuals in the placebo control group had detectable neutralizing activity (8/35), and those who were positive had a low titer resulting in a 50% inhibitory dilution (ID_50_) geometric mean titer (GMT) of 1:5.9 (**Figure 1A**). NDV-HXP-S ID_50_ titers ranged from 1:73.4 (1μg) to 1:231.1 (10μg). A difference between adjuvanted and nonadjuvanted formulations was only found for the 3μg dose with 1:123.0 and 1:101.1 GMT, respectively. BNT162b2 recipient sera had a GMT of 1:146.8, whereas the HCS had a GMT of 1:68.0. While these differences were large, no statistical significance between the groups could be established in a one-way ANOVA when corrected for multiple comparisons due to small group size. Similar activity was detected against the B.1.617.2 (Delta) and B.1.351 (Beta) variants, although – as expected – with reduced titers (**Figure 1B and C**).

**Figure 1:**
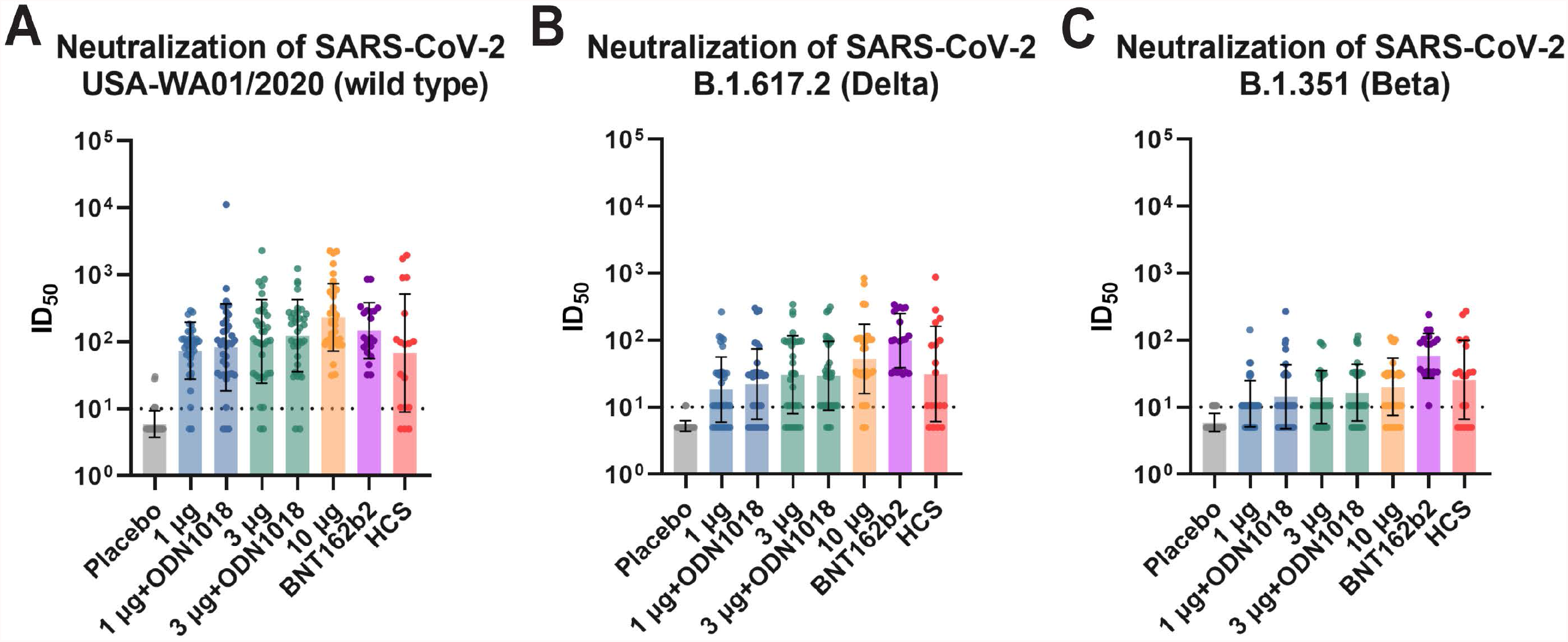
Neutralizing activity of vaccinee and convalescent sera against wild type SARS-CoV-2 and the Delta and Beta variants. Neutralization was measured against wild type SARS-CoV-2 strain USA-WA01/2020 (**A**), a Delta (B.1.617.2) isolate (**B**) and a Beta (B.1.351) isolate (**C**) in a microneutralization assay with authentic SARS-CoV-2. For vaccine groups N=35, 20 individuals were included in the BNT162b2 group and sera from 18 individuals were included in the HCS group. The exception is the 3μg group where only 34 samples were tested in **B** and only 31 in **C**, the 1ug and 1 ug + ODN1018 groups in **C** where n=34 and the placebo group in **C** where n=33 due to a lack of sample volume. Bars show geometric mean titer (GMT), error bars indicate standard deviation of the GMT. The dotted line indicates the limit of detection; values below the limit of detection were assigned a value of half of the limit of detection.

### Binding activity of sera from NDV-HXP-S vaccinees is lower than that of sera from BNT162b2 vaccinees

Next, we assessed binding to wild type spike protein using the commercial SeroKlir Kantaro Semi-Quantitative SARS-CoV-2 IgG Antibody Kit (*22*). Interestingly, binding antibody titers were much lower in this binding assay for the NDV-HXP-S vaccinees as compared to the BNT162b2 vaccinees (**Figure 2A**). Furthermore, the 10μg group was on par with the HCS group while other vaccine groups were slightly lower than the convalescents. To confirm these findings, we also tested binding to wild type spike in a research grade enzyme linked immunosorbent assay (ELISA) (*23*) (**Figure 2B**). Similar to what we observed in the Kantaro assay, the NDV-HXP-S vaccine induced decreased binding titers against wild type full-length spike compared to the BNT162b2 vaccine (**Figure 2B**). We also tested binding to an extensive array of variant spike proteins (**Figure 2B**). While binding was maintained across all variants tested, this pattern of lower binding activity of sera from NDV-HXP-S vaccinees was seen across the board.

**Figure 2:**
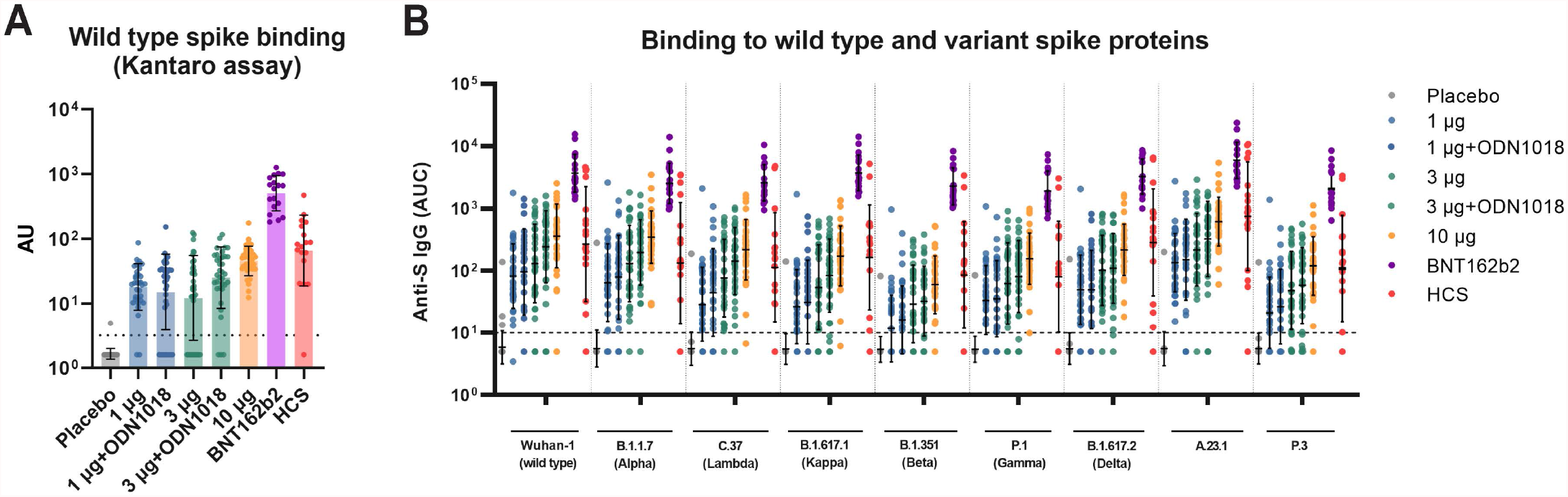
Binding activity of vaccinee and convalescent sera against wild type and variant spike proteins. In **A**, binding to wild type spike protein was assessed using the SeroKlir Kantaro Semi-Quantitative SARS-CoV-2 IgG Antibody Kit. In **B**, a research grade ELISA was used to assess binding to wild type as well as variant spike proteins. For vaccine groups N=35, 20 individuals were included in the BNT162b2 group and sera from 18 individuals were included in the HCS group. The exception was the BNT162b2 group for which N was 18 in **A**. Bars show geometric mean titers (GMT), error bars indicate standard deviation of the GMT. The dotted line indicates the limit of detection; values below the limit of detection were assigned a value of half of the limit of detection.

### The NDV-HXP-S vaccine induces an antibody response that consists of a high proportion of neutralizing antibodies

Since the neutralizing antibody titers of sera from NDV-HXP-S and BNT162b2 vaccinees were similar, and binding titers were much lower for NDV-HXP-S, we decided to determine ratios between binding and neutralizing titers (binding titers taken from **Figure 2B**, neutralizing titers taken from **Figure 1A)**. As observed before (*10, 15*), the ratio of binding to neutralizing antibodies was much better (lower, indicating a higher proportion of neutralizing antibodies) in convalescent individuals as compared to individuals vaccinated with BNT162b2 (**Figure 3**). Surprisingly, the ratio of binding to neutralizing antibodies in NDV-HXP-S vaccinated individuals was similar to or even better than that of convalescent individuals, suggesting that a large proportion of the antibodies induced by this vaccine had neutralizing activity. To confirm these findings, randomly selected samples from the complete sample set were sent to an independent laboratory (the Suthar laboratory at Emory University) for validation of our findings. The laboratory was asked to measure binding and neutralization activities but the provided samples were blinded, and the reason for running the samples was not disclosed. The neutralization assay used in this second laboratory consisted of a focus reduction neutralization assay (FRNT), and the binding assay used was based on the MesoScale Discovery platform, in contrast to the microneutralization assay and ELISA used at Mount Sinai. While the ratios themselves were different (as to be expected due to the different methods used), the difference between convalescent sera and sera from mRNA vaccinated individuals was maintained (**Figure 3B**). Sera from NDV-HXP-S vaccinated individuals again showed ratios similar to sera from convalescent individuals. We also assessed these ratios for the Delta and Beta variants since both specific neutralizing activity and binding to the respective variant spikes were available. The pattern seen with wild type SARS-CoV-2 was also observed with these two viral variants (**Figure 3C** and **D**). Importantly, the ratios of binding to neutralizing activities in sera from BNT162b2 vaccinated individuals were significantly different from ratios measured in sera from NDV-HXP-S vaccinated or from convalescent individuals.

**Figure 3:**
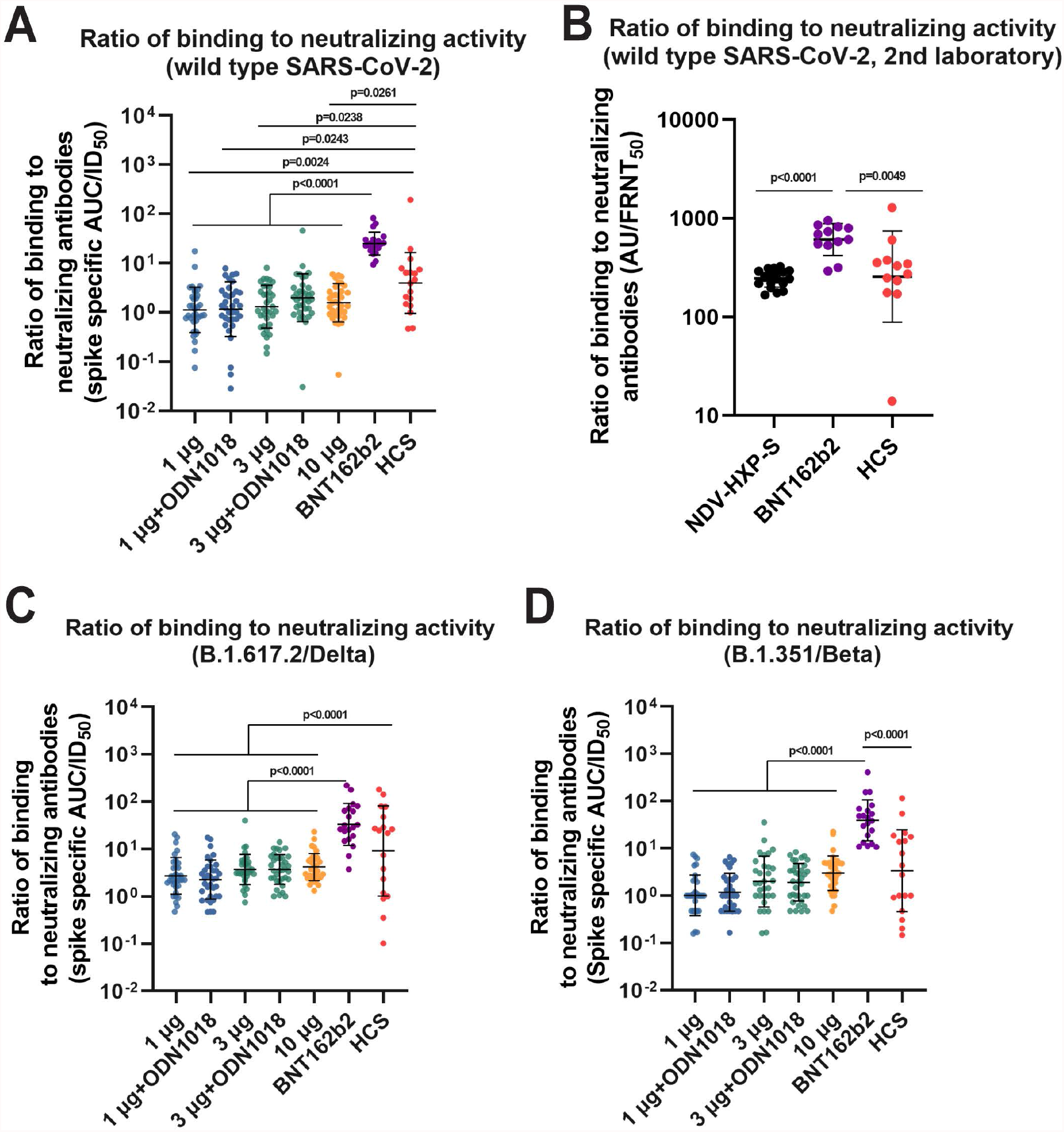
Ratios of binding to neutralizing activity. **A** shows ratios between binding and neutralizing activity for wild type SARS-CoV-2. Binding was analyzed using a research grade ELISA (Figure 2B) and neutralization was assessed using a microneutralization assay (Figure 1A). **B** shows a subset of samples analyzed blindly in an independent laboratory using the MesoScale Discovery platform for binding and an FRNT_50_ assay for neutralization. **C** shows ratios for Delta using Delta binding and neutralization data (from Figures 2B and 1B) and **D** shows ratios for Beta using Beta binding and neutralization data (from Figures 2B and 1C). Geometric mean ratios are shown; error bars indicate standard deviation of the GMR. For vaccine groups N=35, 20 individuals were included in the BNT162b2 group and sera from 18 individuals were included in the HCS group. The exception was **B** for which a subset of 19 NDV-HXP-S samples was tested and the N for BNT162b2 and HCS was 12. In addition, in **C** the N for the 3 ug group is 34 and in **D** the N for the 1ug and 1ug+ODN1018 is 34 and for the 3 ug group it is 31 due to the lack of neutralization data. Statistical analysis was performed using a one-way ANOVA corrected for multiple comparisons. Statistical significant differences are indicated in the figure.

### NDV-HXP-S drives an RBD focused immune response with little NTD and S2 antibodies induced

To determine which domains of the spike protein are targeted in convalescent, BNT162b2 mRNA vaccinated, and NDV-HXP-S vaccinated individuals we then performed ELISAs against recombinant RBD, NTD, and S2 proteins. Sera from all NDV-HXP-S vaccine regimens as well as BNT162b2 vaccination and natural infection, displayed strong RBD titers, albeit at different magnitudes (**Figure 4A**). NTD antibodies were predominantly only present at high titers in BNT162b2 vaccinated individuals (**Figure 4B**). Finally, S2 antibodies were strongly induced by natural infection and by BNT162b2 vaccination but to a much lower degree by NDV-HXP-S vaccination (**Figure 4C**). These data suggest that the strong neutralizing activity after NDV-HXP-S vaccination is likely driven by an RBD-focused response.

**Figure 4:**
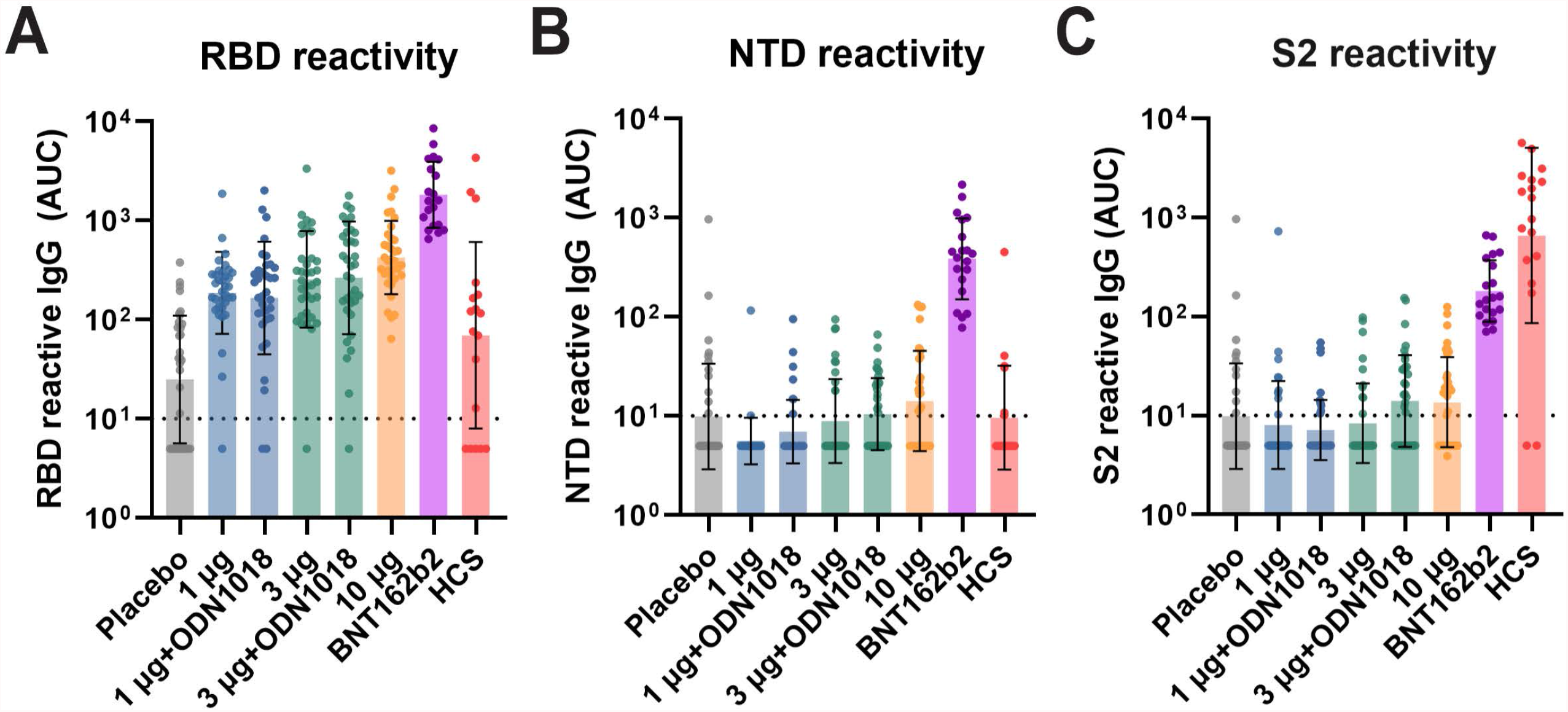
Binding activity of vaccine and convalescent sera to RBD, NTD, and S2. **A** shows binding of the serum panel to RBD, **B** shows binding to NTD and **C** shows binding to recombinant S2 protein. For vaccine groups N=35, 20 individuals were included in the BNT162b2 group and sera from 18 individuals were included in the HCS group. Bars show geometric mean titers (GMT), error bars indicate standard deviation of the GMT. The dotted line indicates the limit of detection, values below the limit of detection were assigned a value of half of the limit of detection.

## Discussion

During the COVID-19 pandemic, global vaccine distribution and vaccine equity were, and continue to be, suboptimal. Locally produced vaccines can increase vaccine access and vaccine independence, especially for LMICs. The NDV-HXP-S vaccine is designed to help close this gap since it can be economically produced in influenza vaccine manufacturing plants that are located in LMICs. Moreover, it can be stored and distributed without the need for freezers and incorporates an advanced HexaPro antigen design compared to most other COVID-19 vaccines on the market (*3*). The NDV-HXP-S vaccine development program also provides a vaccine platform and model that can be used for optimal pandemic preparedness and response in LMICs in the future. Importantly, here we show that an inactivated version of the NDV-HXP-S vaccine is capable of inducing neutralizing antibody titers in humans that are comparable to titers induced by the BNT162b2 mRNA vaccine (Pfizer). Interestingly, while the induced neutralizing activity is comparable with mRNA vaccination, the binding antibody titers are much lower. This results in a response dominated by neutralizing antibodies, as observed when comparing ratios of binding to neutralizing antibody titers. It has been shown previously by our team and others, that mRNA vaccination – while inducing very high and protective neutralizing antibody titers – also induces a large quantity of non-neutralizing antibodies that, on a monoclonal level, target RBD, NTD, and S2 (*10, 15*). In fact, while neutralizing titers after natural infection are usually lower than after mRNA vaccination, the ratio of binding to neutralizing antibodies is more favorable. Here, we observe comparable ratios for convalescent individuals and NDV-HXP-S vaccinated individuals, while the ratios for mRNA vaccinees are significantly different. We confirmed these findings by using two different assay formats conducted in independent research laboratories for measuring both binding and neutralization activity. When analyzing the polyclonal response for binding to RBD, NTD, and S2, we found that mRNA vaccination induces strong immune responses against all three targets; natural infection mostly targets RBD and S2, while NDV-HXP-S targets almost exclusively the RBD. This explains the high proportion of neutralizing antibodies since most epitopes targeted by neutralizing antibodies are located in the RBD. There could be two reasons for this very focused immune response. First, presentation of the spike on the NDV particle could limit access of B-cell receptors to the extreme membrane distal part of the spike protein – which is the RBD. Akin to a dense forest of mature trees, the NDV-HXP-S particles are likely densely packed with NDV hemagglutinin-neuraminidase (HN), NDV fusion protein (F), and SARS-CoV-2 spike protein. It is easy to imagine, that B-cell receptors, which are also surrounded by many other membrane proteins on the surface of B-cells, could be sterically hindered from reaching the membrane proximal S2 domain or the NTD, which is also located closer to the membrane than the RBD. The second hypothesis is, that the HexaPro antigen design (*3*), which arrests the spike protein in the pre-fusion conformation, leads to a refocusing of the antibody response to the RBD. Many S2 epitopes may only be accessible in the post-fusion conformation and may be exposed in wild type spike or the metastable (2P) spike included in mRNA vaccines. These S2 epitopes may distract the immune responses from the RBD. A pre-fusion stabilized spike like the HexaPro may not display these epitopes, hence focusing most of the immune response to the RBD. Of course, the effect observed could also be caused by a combination of both mechanisms.

Our study has several limitations. We include a limited number of samples, and our dataset comes from a comparison of imperfectly matched groups from a clinical trial in Thailand and an observational cohort study in New York City. Strengths of the study include the use of authentic SARS-CoV-2 for neutralization assays and that the findings could be replicated with different methods in a different, blinded, and independent laboratory.

In summary, we show that a vaccine candidate which can be produced locally in LMICs at low cost induces neutralizing antibody titers to SARS-CoV-2 comparable to those observed in cohorts having received mRNA-based COVID-19 vaccines. The NDV-HXP-S vaccine candidate induces a strong RBD focused immune response resulting in a high proportion of neutralizing antibodies, which are associated with protection from infection and severe disease (*20, 24, 25*).

## Methods

### Human Serum Samples

Sera collected from the NDV-HXP-S clinical trial in Thailand (placebo, 1μg, 1 μg + ODN1018, 3μg, 3μg + ODN1018, and 10μg, n=35 samples per group) were used in this study. Characteristics of the clinical trial samples are indicated in **Table 1**. Additional detail can be found in the published interim report (*9*). In addition, we selected convalescent (n=18) and post-vaccine sera (n=20) that were collected from participants in two longitudinal observational studies. The BNT162b2 vaccinees were selected from the PARIS (Protection Associated with Rapid Immunity to SARS-CoV-2) cohort (*21*), while the convalescent serum samples were selected from PARIS as well as from our observational virus infection cohort to best match the demographics of the vaccine trial participants. The PARIS cohort follows health care workers of the Mount Sinai Health System longitudinally since April 2020 while the observational virus infection cohort is open to anyone willing to participate. These studies were reviewed and approved by the Mount Sinai Hospital Institutional Review Board (IRB-20-03374,IRB-16-00791). All participants signed written consent forms prior to sample and data collection. All participants provided permission for sample banking and sharing. All samples were stripped of identifying information before distribution to the participating laboratories.

### Cells

Vero.E6 cells were cultured in Dulbecco’s modified Eagles medium (DMEM) containing 10% heat-inactivated fetal bovine serum (FBS), supplemented with 100 U/ml penicillin and 100 μg/ml streptomycin (Gibco).

### Recombinant variant RBD, NTD, S2 and spike proteins

The recombinant RBD and spike proteins used in Figure 2B (Wuhan-1, B.1.1.7) and Figure 4A were produced using Expi293F cells (Life Technologies). The sequences for the proteins were cloned into a mammalian expression vector, pCAGGS, as previously described and proteins were purified after transient transfections with each respective plasmid (*23, 26*). Six hundred million Expi293F cells were transfected using the ExpiFectamine 293 Transfection Kit and purified DNA. Supernatants were collected on day four post-transfection, centrifuged at 4,000 g for 20 minutes, and filtered using a 0.22 μm filter. Ni-nitrilotriacetic acid (Ni-NTA) agarose (Qiagen) was used to purify the proteins by gravity flow. The proteins were eluted as previously described. Buffer exchange was performed using Amicon centrifugal units (EMD Millipore), and all recombinant proteins were re-suspended in phosphate-buffered saline (PBS). Proteins were run on sodium dodecyl sulphate (SDS) polyacrylamide gels (5–20% gradient; Bio-Rad) to check for purity. NTD (Catalogue # 40591-V49H) and S2 (Catalogue #40590-V08B) recombinant proteins were acquired from Sino Biological.

Trimeric spike proteins in Figure 2B (all except Wuhan-1 and B.1.1.7) were produced as previously described (*27*). Briefly, residues 1-1208 of the spike protein (Wuhan-1 strain numbering) were codon optimized with proline substitutions at residues 986 and 987, the furin cleavage site modified to “GSAS”, and a T4 fibritin trimerization motif and a 8X HIS tag were added on the C-terminus and the construct was cloned into pCDNA3.4. 293F cells were transfected with plasmid using PEIMax in FreeStyle 293 Expression Medium (Fisher) and cultured for three days at 32°C, 5% CO_2_. Trimers were purified by Ni-NTA ion exchange chromatography followed by size exclusion chromatography on a HiLoad 16/60 Superdex 200 prep grade size exclusion column (GE Healthcare), and then were buffer exchanged into HEPES (4-(2-hydroxyethyl)-1-piperazineethanesulfonic acid)-buffered-saline with 10mM ethylenediaminetetraacetic acid (EDTA). Antigenicity was verified by ELISA and Octet bilayer interferometry. To create variants of concern, the designated amino acid substitutions corresponding to each variant were introduced into the Wuhan-1 sequence and purified as described above.

### *In-house* enzyme-linked immunosorbent assay (ELISA)

Antibody titers in sera were assessed using a research-grade ELISA (*23*) with recombinant versions of the RBD, NTD, S2, and full-length spike of wild type SARS-CoV-2, as well as the spike from B.1.1.7 (Alpha), C.37 (Lambda), B.1.617.1 (Kappa), B.1.351 (Beta), P.1 (Gamma), B.1.617.2 (Delta), A.23.1 and P.3. Briefly, 96-well microtiter plates (Corning) were coated with 50 μl/well of the corresponding recombinant protein (2 μg/ml) overnight at 4 °C. After three washes with phosphate-buffered saline (PBS) supplemented with 0.1% Tween-20 (PBS-T) using an automatic plate washer (BioTek 405TS microplate washer), plates were blocked with PBS-T containing 3% milk powder (American Bio) for one hour at room temperature (RT). Blocking solution was removed and initial dilutions (1:100) of heat-inactivated sera (in PBS-T 1%-milk powder) were added to the plates, followed by 2-fold serial dilutions and a two hour incubation. After three washes with PBS-T, 50 μl/well of the pre-diluted secondary anti-human IgG (Fab-specific) horseradish peroxidase antibody (produced in goat; Sigma-Aldrich) diluted 1:3,000 in PBS-T containing 1% milk powder were added and plates were incubated for one hour incubation at RT. After three washes with PBS-T, the substrate o-phenylenediamine dihydrochloride (Sigmafast OPD) was added (100 μl/well) for 10min, followed by an addition of 50 μl/well of 3 M hydrochloric acid (Thermo Fisher) to stop the reaction. Optical density was measured at a wavelength of 490 nm using a plate reader (BioTek, SYNERGY H1 microplate reader). Area under the curve (AUC) values were calculated and plotted using Prism 9 software (GraphPad).

### Kantaro enzyme linked immunosorbent assay (ELISA)

Antibody testing with the commercial COVID-SeroKlir Kantaro Semi-Quantitative SARS-CoV-2 IgG Antibody Kit (Kantaro Biosciences, R&D Systems® Catalog Number COV219) was performed as previously described (*22*). This assay has an approximate 99% positive percent agreement and 99% negative percent agreement in PCR+ subjects 15 days post-symptom onset (https://resources.rndsystems.com/pdfs/datasheets/cov219.pdf?v=20210525&_ga=2.12000950.307497989.1621962942-1278575996.1621962942). All reagents and microplates are included with the commercial kit. Briefly, for qualitative RBD ELISAs, samples were diluted in sample buffer (1:100) using 96-well microtiter plates, and 100μl/well of pre-diluted samples were transferred to the RBD pre-coated microplates. Positive and negative controls were added to every plate. Samples were incubated for 2 hours at room temperature. Serum dilutions were removed and plates were washed three times with the included washing buffer. RBD conjugate was diluted in conjugate buffer and 100μl/well were added to the plates for 1 hour. Conjugate was removed and plates were washed three times with wash buffer. To develop the colorimetric reaction, the substrate solution was added (100μl/well) for 20min. 100μl/well of stop solution were added, and plates were read at an optical density (OD) of 450nm and at an OD of 570nm for wavelength correction. As per the manufacturer’s instructions, the cutoff index (CI) was calculated by dividing the corrected OD of the clinical sample by the corrected OD of RBD positive control. Samples with a CI above 0.7 were considered as presumptive positives and were further tested in the confirmatory quantitative ELISA based on the full-length recombinant spike protein.

For the confirmatory quantitative spike ELISA samples were pre-diluted to 1:200 in sample buffer. Sample dilutions were added in duplicate to the pre-coated microplates. Low, medium, and high controls, and spike calibrators used to generate a standard curve, were added to every microtiter plate. After 2 hours of incubation at room temperature, the remaining steps of the ELISA were performed as described above. Data were analyzed using GraphPad Prism 9. The concentration of spike-reactive antibodies was calculated using a four parameter logistic (4-PL) curve-fit. Samples exceeding the range of the standard curve were further diluted and re-tested. Only samples positive in both steps of the assay were considered positive.

### SARS-CoV-2 multi-cycle microneutralization assay

All procedures were performed in the Biosafety Level 3 (BSL-3) facility at the Icahn School of Medicine at Mount Sinai following standard safety guidelines. Vero.E6 cells were seeded in 96-well high binding cell culture plates (Costar) at a density of 20,000 cells/well in complete Dulbecco’s modified Eagle medium (cDMEM) a day before infection. Heat inactivated sera (56°C for 1 hour) were serially diluted (3-fold) in minimum essential media (MEM; Gibco) supplemented with 2mM L-glutamine (Gibco), 0.1% sodium bicarbonate (w/v, HyClone), 10mM 4-(2-hydroxyethyl)-1-piperazineethanesulfonic acid (HEPES; Gibco), 100U/ml penicillin, 100 μg/ml streptomycin (Gibco) and 0.2% bovine serum albumin (MP Biomedicals) starting at a 1:10 dilution. Remdesivir (Medkoo Bioscience inc.) was included as a control to monitor assay variation. Serially diluted sera were incubated with 1,000 tissue culture infectious dose 50 (TCID_50_) of wild type USA-WA1/2020 SARS-CoV-2, B.1.617.2 (Delta) or B.1.351 (Beta) virus isolates for one hour at RT, followed by the transfer of 120μl of the virus-sera mix to Vero.E6 plates. Infection was left to proceed for one hour at 37°C, followed by removal of the inoculum. 100μl/well of the infection media supplemented with 2% fetal bovine serum (FBS; Gibco) and 100μl/well of antibody dilutions were added to the cells. Plates were incubated for 48 hours at 37°C and cell monolayers were fixed with 200μl/well of a 10% formaldehyde solution overnight at 4°C. After removal of the formaldehyde solution, and washing with PBS (pH 7.4) (Gibco), cells were permeabilized by adding 150μl/well of PBS, 0.1% Triton X-100 (Fisher Bioreagents) for 15 min at RT to allow staining of the nucleoprotein (NP). Permeabilization solution was removed and plates were blocked with PBS 3% BSA for 1 hour at RT. The biotinylated mAb 1C7C7, a mouse anti-SARS nucleoprotein monoclonal antibody generated at the Center for Therapeutic Antibody Development at the Icahn School of Medicine at Mount Sinai ISMMS (Millipore Sigma), was used for NP staining at a concentration of 1μg/ml in PBS, 1% BSA. After a 1 hour incubation at RT, cells were washed with 200μl/well of PBS twice and 100μl/well of HRP-conjugated streptavidin (Thermo Fisher Scientific) diluted in PBS, 1% BSA wasadded at a 1:2,000 dilution. Following a 1 hour incubation at RT, cells were washed twice with PBS, and 100μl/well of Sigmafast OPD were added for 10min at RT. Addition of 50μl/well of a 3M HCl solution (Thermo Fisher Scientific) allowed to stop the reaction. The optical density (OD) was measured (490 nm) using a microplate reader (Synergy H1; Biotek). All the analyses were performed using Prism 7 software (GraphPad). A nonlinear regression curve fit analysis was performed to calculate the inhibitory dilution 50% (ID_50_).

### Focus reduction neutralization test

FRNT assays were performed as previously described (*28-30*). Briefly, samples were diluted at 3-fold in 8 serial dilutions using DMEM (VWR, #45000-304) in duplicates with an initial dilution of 1:10 in a total volume of 60 μl. Serially diluted samples were incubated with an equal volume of icSARS-CoV-2 (100-200 foci per well based on the target cell) at 37° C for 45 minutes in a round-bottomed 96-well culture plate. The antibody-virus mixture was then added to Vero.E6-TMPRSS2 cells and incubated at 37°C for 1 hour. Post-incubation, the antibody-virus mixture was removed and 100 μl of pre-warmed 0.85% methylcellulose (Sigma-Aldrich, #M0512-250G) overlay was added to each well. Plates were incubated at 37° C for 18 hours and the methylcellulose overlay was removed and washed six times with PBS. Cells were fixed with 2% paraformaldehyde in PBS for 30 minutes. Following fixation, plates were washed twice with PBS and permeabilization buffer (0.1% BSA [VWR, #0332], saponin [Sigma, 47036-250G-F] in PBS) was added to permeabilized cells for at least 20 minutes. Cells were incubated with an anti-SARS-CoV spike primary antibody directly conjugated to Alexafluor-647 (CR3022-AF647) for up to 4 hours at room temperature. Cells were washed three times in PBS and foci were visualized on a CTL Analyzer. Antibody neutralization was quantified by counting the number of foci for each sample using the Viridot program (*31*). The neutralization titers were calculated as follows: 1 - (ratio of the mean number of foci in the presence of sera and foci at the highest dilution of respective sera sample). Each specimen was tested in duplicate. The FRNT_50_ titers were interpolated using a 4-parameter nonlinear regression in GraphPad Prism 9.2.0. Samples that do not neutralize at the limit of detection at 50% were plotted at 10 for geometric mean and fold-change calculations.

### Multiplex Immunoassay

Serum samples were tested for their IgG binding against the SARS-CoV-2 Wuhan-1 strain using an electro chemiluminescent-based multiplex immunoassay provided by Mesoscale Discovery (MSD-ELICA). The experiment was performed according to the manufacturer’s instructions. The COVID-19 Coronavirus Panel 1 (Catalog No. K15362U) was used for measuring spike antibody binding titers. Plates were pre-coated with the antigens. Briefly, blocking was performed for minimum of 30 min, with 150 μL per well of MSD Blocker A. To assess binding, samples were diluted 1:5000. 50 μL of each sample and Reference Standard dilution were added to the plates in duplicate and incubated for 2 hours. Following this, 50 μL per well of 1X MSD SULFO-TAG Anti-Human IgG detection antibodies were added and incubated for 1 hour. Following the detection reagent step, 150 μL per well of MSD Gold Read Buffer B was added to each plate immediately prior to reading on an MSD plate reader (MESO QuickPlex SQ 120). Plates were washed three times with 300 μL PBS/0.05% Tween 20 between each step. At every incubation step, plates were kept at RT and shaking with at a speed of 700 rpm. Data was analyzed using Discovery Workbench software. The antibody concentration in arbitrary units (AU) was calculated relative to the provided Reference Standard.

### Statistics

A one-way ANOVA with correction for multiple comparisons test was used to compare the neutralization titers and ratios. In some cases, two groups were compared using a Student’s t-test. Statistical analyses were performed using Prism 9 software (GraphPad).

## Data Availability

All data produced in the present study are available upon reasonable request to the authors.

## Data availability statement

All data produced in the present study are available upon reasonable request to the authors.

## Acknowledgments

We thank the study participants of the vaccine trial, the PARIS cohort and our longitudinal observational cohort for their generosity and willingness to help advance our knowledge on SARS-CoV-2 immune responses. We thank Dr. Randy A. Albrecht for oversight of the conventional BSL3 biocontainment facility at Mount Sinai, which makes our work with live SARS-CoV-2 possible. We are also grateful for Mount Sinai’s leadership during the COVID-19 pandemic. We want to especially thank Drs. Dennis Charney, David Reich, and Kenneth Davis for their support. We also would like to thank the teams at PATH, Mahidol University and the Government Pharmaceutical Organization for their support.

This work is part of the PARIS/SPARTA studies funded by the NIAID Collaborative Influenza Vaccine Innovation Centers (CIVIC) contract 75N93019C00051. In addition, this work was also partially funded by the NIAID Centers of Excellence for Influenza Research and Response (CEIRR) contract and 75N93021C00014 and 75N93021C00017 and by anonymous donations to Mount Sinai. Work on NDV-HXP-S vaccines was also supported by the Bill & Melinda Gates Foundation and by institutional funding from the Icahn School of Medicine at Mount Sinai. PATH funded the shipment of samples from Nexelis to Mount Sinai. The main NDV-HXP-S project in Thailand is funded by the National Vaccine Institute, Government Pharmaceutical Organization.

## References

1. F. Krammer, SARS-CoV-2 vaccines in development. Nature 586, 516–527 (2020).

2. W. Sun et al., A Newcastle disease virus expressing a stabilized spike protein of SARS-CoV-2 induces protective immune responses. Nat Commun 12, 6197 (2021).

3. C. L. Hsieh et al., Structure-based design of prefusion-stabilized SARS-CoV-2 spikes. Science, (2020).

4. E. Sparrow et al., Global production capacity of seasonal and pandemic influenza vaccines in 2019. Vaccine 39, 512–520 (2021).

5. W. Sun et al., A Newcastle Disease Virus (NDV) Expressing a Membrane-Anchored Spike as a Cost-Effective Inactivated SARS-CoV-2 Vaccine. Vaccines (Basel) 8, (2020).

6. W. Sun et al., Newcastle disease virus (NDV) expressing the spike protein of SARS-CoV-2 as a live virus vaccine candidate. EBioMedicine 62, 103132 (2020).

7. J. Tcheou et al., Safety and Immunogenicity Analysis of a Newcastle Disease Virus (NDV-HXP-S) Expressing the Spike Protein of SARS-CoV-2 in Sprague Dawley Rats. Front Immunol 12, 791764 (2021).

8. J. H. Lara-Puente et al., Safety and Immunogenicity of a Newcastle Disease Virus Vector-Based SARS-CoV-2 Vaccine Candidate, AVX/COVID-12-HEXAPRO (Patria), in Pigs. mBio, e0190821 (2021).

9. P. Pitisuttithum et al., Safety and Immunogenicity of an Inactivated Recombinant Newcastle Disease Virus Vaccine Expressing SARS-CoV-2 Spike: Interim Results of a Randomised, Placebo-Controlled, Phase 1/2 Trial. medRxiv, (2021).

10. F. Amanat et al., SARS-CoV-2 mRNA vaccination induces functionally diverse antibodies to NTD, RBD, and S2. Cell 184, 3936-3948.e3910 (2021).

11. G. Cerutti et al., Potent SARS-CoV-2 neutralizing antibodies directed against spike N-terminal domain target a single supersite. Cell Host Microbe 29, 819-833.e817 (2021).

12. M. F. Jennewein et al., Isolation and characterization of cross-neutralizing coronavirus antibodies from COVID-19+ subjects. Cell Rep 36, 109353 (2021).

13. M. McCallum et al., N-terminal domain antigenic mapping reveals a site of vulnerability for SARS-CoV-2. Cell, (2021).

14. C. Gaebler et al., Evolution of Antibody Immunity to SARS-CoV-2. bioRxiv, (2020).

15. A. Cho et al., Anti-SARS-CoV-2 receptor-binding domain antibody evolution after mRNA vaccination. Nature 600, 517–522 (2021).

16. D. Pinto et al., Broad betacoronavirus neutralization by a stem helix-specific human antibody. Science 373, 1109–1116 (2021).

17. F. Amanat et al., SARS-CoV-2 mRNA vaccination induces functionally diverse antibodies to NTD, RBD, and S2. Cell, (2021).

18. J.M. Carreño et al., Evidence for retained spike-binding and neutralizing activity against emerging SARS-CoV-2 variants in serum of COVID-19 mRNA vaccine recipients. EBioMedicine 73, 103626 (2021).

19. J.M. Carreño et al., Activity of convalescent and vaccine serum against SARS-CoV-2 Omicron. Nature, (2021).

20. K. A. Earle et al., Evidence for antibody as a protective correlate for COVID-19 vaccines. Vaccine, (2021).

21. F. Krammer et al., Antibody Responses in Seropositive Persons after a Single Dose of SARS-CoV-2 mRNA Vaccine. N Engl J Med 384, 1372–1374 (2021).

22. J.M. Carreño et al., Longitudinal analysis of severe acute respiratory syndrome coronavirus 2 seroprevalence using multiple serology platforms. iScience 24, 102937 (2021).

23. D. Stadlbauer et al., SARS-CoV-2 Seroconversion in Humans: A Detailed Protocol for a Serological Assay, Antigen Production, and Test Setup. Curr Protoc Microbiol 57, e100 (2020).

24. P. B. Gilbert et al., Immune correlates analysis of the mRNA-1273 COVID-19 vaccine efficacy clinical trial. Science, eab3435 (2021).

25. D. S. Khoury et al., Neutralizing antibody levels are highly predictive of immune protection from symptomatic SARS-CoV-2 infection. Nat Med, (2021).

26. F. Amanat et al., A serological assay to detect SARS-CoV-2 seroconversion in humans. Nat Med, (2020).

27. W. E. Harrington et al., Rapid decline of neutralizing antibodies is associated with decay of IgM in adults recovered from mild COVID-19. Cell Rep Med 2, 100253 (2021).

28. A. Vanderheiden et al., Development of a Rapid Focus Reduction Neutralization Test Assay for Measuring SARS-CoV-2 Neutralizing Antibodies. Curr Protoc Immunol 131, e116 (2020).

29. V. V. Edara et al., Infection- and vaccine-induced antibody binding and neutralization of the B.1.351 SARS-CoV-2 variant. Cell Host Microbe 29, 516–521 e513 (2021).

30. V. V. Edara et al., Infection and Vaccine-Induced Neutralizing-Antibody Responses to the SARS-CoV-2 B.1.617 Variants. N Engl J Med 385, 664–666 (2021).

31. L. C. Katzelnick et al., Viridot: An automated virus plaque (immunofocus) counter for the measurement of serological neutralizing responses with application to dengue virus. PLoS Negl Trop Dis 12, e0006862 (2018).

